# Deprivation effect on COVID-19 cases incidence and severity: a geo-epidemiological study in PACA region, France

**DOI:** 10.1101/2023.04.18.23288723

**Authors:** Guillaume Gaubert, Steve Nauleau, Florian Franke, Stanislas Rebaudet, Emilie Mosnier, Jordi Landier, Pascal Chaud, Philippe Malfait, Stéphanie Vandentorren, Michael Huart, Alaa Ramdani, Marc-Karim Bendiane, Fabrice Danjou, Jean Gaudart

## Abstract

**Introduction:** The spread of the COVID-19 pandemic, and its severity, is spatially heterogenous. At the individual level, the socioeconomic status (SES) profile is known to be associated with COVID-19 incidence and severity. The aim of this geo epidemiological study was to investigate the link between SES profile and potential confounders, and COVID-19 incidence and hospitalization rates, at a fine geographical scale.

**Methods:** We analyzed COVID-19 incidence and severity during two epidemic waves between September 2020 and June 2021, in Provence Alpes Côtes d’Azur, a 5 million inhabitants’ French region. The region is divided into sub-municipal areas that we have classified according to their SES profile. We then conducted a spatial analysis of COVID-19 indicators depending on SES profile, age structure, and health services provision. This analysis considered spatial autocorrelation between areas.

**Results:** COVID-19 incidence rates in more deprived areas were similar to those in wealthiest ones. Hospitalization rates of COVID-19 cases in conventional care units were greater in more deprived vs wealthiest areas: Standardized Incidence Ratio (SIR) were respectively 1.34 [95% confidence interval 1.18 - 1.52] and 1.25 [1.13 - 1.38] depending on the epidemic wave. This gap was even greater regarding hospitalization rates of cases in critical care units: SIR = 1.64 [1.30 - 2.07] then 1.33 [1.14 - 1.55] depending on the epidemic wave. Hospitalization rates of COVID-19 cases in conventional care units were also greater in areas with high proportion of elderly people vs young people: SIR respectively 1.24 [1.11 - 1.38] and 1.22 [1.13 - 1.32] depending on the wave.

**Conclusion:** Considering age structure and health services provision, a deprived SES profile is associated to a greater COVID-19 severity in terms of hospitals admissions, in conventional care units and in critical care units. This result implies targeting risk prevention efforts on these areas in pandemic situations, and highlights the need to develop access to healthcare to deprived populations in anticipation of periods of crisis.

**Key messages:** **What is already known on this topic** - Socioeconomic status is associated to COVID-19 incidence and severity, at an individual scale or at a large spatial scale.

**What this study adds** - We showed the positive relationship between deprivation and COVID-19 incidence and hospitalization rates at a fine sub-municipal geographical scale. We considered confusion factors like demographic structure and health services provision.

**How this study might affect research, practice or policy** - These findings may help predict at a fine scale where the impact will be most severe in pandemic situations and make it possible to target risk prevention efforts on these areas.

## Introduction

The spread of the COVID-19 pandemic in France was heterogenous across administrative departments^1^, or neighborhoods^2, 3^. This has been described for the severity too, estimated by the in-hospital COVID-19 incidence and mortality^1, 4^.

Studies have been done on the area-level spatial differences associated with the spread of the pandemic, or the occurrence of death^3, 5^. However, few ones have been conducted on these population risk factors associated with COVID-19 severity^6, 7^, or at a large geographical scale^1^. At the individual level, an older age, male sex or certain chronic conditions such as obesity, diabetes and COPD (Chronic obstructive pulmonary disease)^8, 9^ as well as social position^10^ and ethnicity^11^ are related with greater severity. At population-level, studies identified X, Y and Z as factors associated with covid-19 spread, and X associated with covid-19 relate deaths. However, studies addressing heterogeneity of severe disease and population-level risk factors remain limited.

In several countries, deprivation was associated to higher COVID-19 death rate^12^ and hospitalization risk^13^. The existence of this link is supported by the one between deprivation and increased comorbidities such as diabetes^14^, obesity^15^ or COPD^16^. The barriers to access to healthcare and the renunciation of healthcare for deprived populations could also play a role, as well as the greater impact of income loss from the formal and informal economy during lockdowns.

In France where universal social and health coverage exists, no population-based studies have assessed the association of socio-economic status (SES) profile of residence area with the COVID-19 incidence and the severity of these cases.

In this geo-epidemiological study, we assessed determinants associated with COVID-19 case incidence rates, conventional units hospitalizations-to-cases ratios, and critical care units hospitalizations-to-cases ratios, at the local scale, between fall 2020 and spring 2021, in the Provence-Alpes-Côte d’Azur (PACA) administrative region, South Eastern France.

## Material and methods

### Study design

This ecological population-based study aimed to assess the impact of social deprivation on COVID-19 on a fine geographical scal. The PACA region is located in south-eastern France with a population of 5 million inhabitants, of which 88% are concentrated in urban areas along the Mediterranean coast^17^. We also considered, as potential determinants, age structure and health services provision.

### Study period

We analysed the periode between 2020-09-01, and 2021-06-01 corresponding to the second and third wave of cases recorded in metropolitan France. The split date between these 2 periods was set at December 15, corresponding to the end of the lockdown started in November 2021, the start of school holidays and end-of-year family reunions, and a low incidence rate.

### Data sources

All COVID-19 cases detected in the population were extracted from the national information system implemented on May 13, 2020 by Sante publique France (the French National Agency of Public Health), called SIDEP (“Système d’Information de Dépistage Populationnel”). SIDEP is a secure platform that records all SARS-CoV-2 RT-PCR and antigen test results from laboratories, hospitals, pharmacies, nurses, and physicians across France, including information on sex, age, and residential address.

Hospital admissions of COVID-19 diagnosed patients were reported by hospital services to Santé publique France, through another national information system called SIVIC (“Système d’Information pour le suivi des VICtimes”). Two types of units were considered: conventional care units and critical care units (including, in France, resuscitation, intensive care and continuing care units).

In France, the socio-demographic characteristics of the population are published by the National Institute of Statistics (INSEE) at the IRIS level (Regrouped Islets for Statistical Information) on its open data website^18^. IRIS correspond to an area with 1,000 to 5,000 inhabitants. To assess the impact of social deprivation, we extracted all the variables available related to the SES profile of the IRIS: population density, proportion of each socio-professional category, proportion of foreigners and immigrants, bachelor’s degree rate, unemployed rate, median income, European Deprivation Index^19^, and proportion of overcrowded homes. Finally, we extracted from INSEE website variables related to the demographics (proportion of different age group [< 18; 18-39; 40-64; ≥ 65]) and health services provision in these IRIS, since we suspected these parameters to be confounding factors.

We selected the four following variables to characterize health services provision: presence of a biological laboratory (usual place for screening tests); presence of a retirement home; number of frontline caregivers (listed in Appendix 5); distance between IRIS centroid and nearest emergency department (in kilometers); and access to general practitioners (measured by the number of possible consultations per person and per year)^20^. This last variable was only available at the municipality-level.

All data sources are presented in a table in the Appendix 6.

### Data aggregation

Data aggregation was performed by the regional health authority, (“Agence Régionale de Santé, Provence Alpes Cote d’Azur”, ARS-PACA) for the cases and hospital admissions, which linked residential address of COVID-19 cases or admitted patients to the corresponding IRIS.

The level of temporal aggregation was the wave as previously defined, according to the day of screening for cases, and the day of admission for hospital data.

### Indicators

To consider the different ways in which COVID-19 affected the population, we chose to study three major indicators: cases, admissions to conventional care units, and admissions to critical care units.

First, the COVID-19 incidence rate was calculated as the ratio of the number of cases reported during the epidemic wave to the number of inhabitants in the area considered, during the same period.

Second, we calculated two severity ratios: the ratio of the number of hospital admissions in conventional units for COVID-19 to the number of detected COVID-19 cases during the same period in the same IRIS; and the same ratio for critical care units (CCU) admissions for COVID-19.

### IRIS selection criteria

PACA region has 2446 IRIS (5.04 million inhabitants). INSEE classifies them into 3 types: habitat, activity, and diverse. We excluded 139 “activity” IRIS (that may describe people working in the IRIS more than people living there), “diverse” IRIS, and “habitat” IRIS with less than 30 inhabitants (as it could lead to a risk of subjects’ identification according to the French regulation) and 177 IRIS with no hospitalization data. Finally, 2130 IRIS (4.92 million inhabitants) were included in our geo-epidemiological analysis (Appendix 1).

### Statistical analysis

We mapped these three indicators for each IRIS and pandemic wave, and we represented their evolution over time. We then estimated the spatial autocorrelation between IRIS for each one by the Moran’s index, using Queen’s criterion of contiguity^21^.

Deprivation is defined relatively to the other members of the community or society^22^. To identify more deprived IRIS, we therefore built SES profiles using hierarchical ascendant classification based on principal component analysis coordinates (HCPC), an unsupervised approach that allowed using all SES variables previously mentioned. This method avoids the curse of dimensionality and collinearity problems^23^. Age structure profiles were similarly defined, using the proportion of each age category per IRIS. All included variables in both classifications are described in Appendix 7.

For each period (second and third wave), the associations between suspected determinants (SES and age structure profiles, health services provision) and the COVID-19 indicators, were analyzed by using generalized additive mixed models (GAMM). We parameterized a negative binomial distribution (NegBin) to consider overdispersion of admissions data. A random effect (RE(city)) was included modeling the municipality-level impact, as the access to general practitioner was available at this scale. To consider spatial correlations, we used a Gaussian kriging smoother^24^ of the geographical coordinates of each IRIS centroid (s(longitude, latitude)). The log transformed number of inhabitants or cases in offset terms allowed us to model incidence and severity ratios (conventional-hospitalizations-to-cases ratios and critical-care-admissions-to-cases ratios), respectively. Final models had the following form:

#### Incidence rate

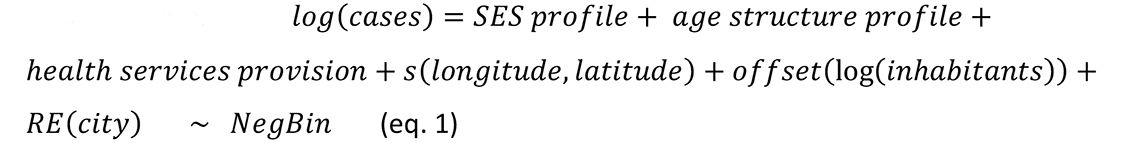

#### Severity ratios

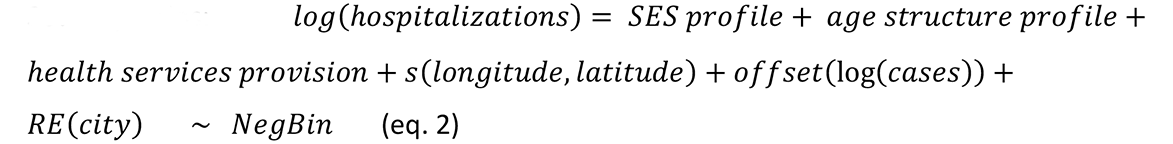

The spatial autocorrelation of the model residuals was assessed by the Moran statistic as a sensitivity analysis to confirm the absence of residual spatial autocorrelation.

All analyses and maps were made using the software program R version 4.0.2 (R Development Core Team, R Foundation for Statistical Computing, Vienna, Austria) with the following main packages: {FactoMineR}^23^, {mgcv}^24^, and {spdep}^25^.

## Results

### COVID-19 indicators description

The second pandemic wave in the PACA region lasted 3.5 months (from 2020-09-01 to 2020- 12-14) and the mean incidence rate of confirmed COVID-19 was 233.2 cases per week per 100,000 inhabitants with a peak at 501.7. The third wave lasted longer (5.5 months, from 2020-12-15 to 2021-06-01) and mean incidence rate was higher at 257.9, but the peak was lower at 366.8 (Fig 1, panel A).

**Figure 1.**
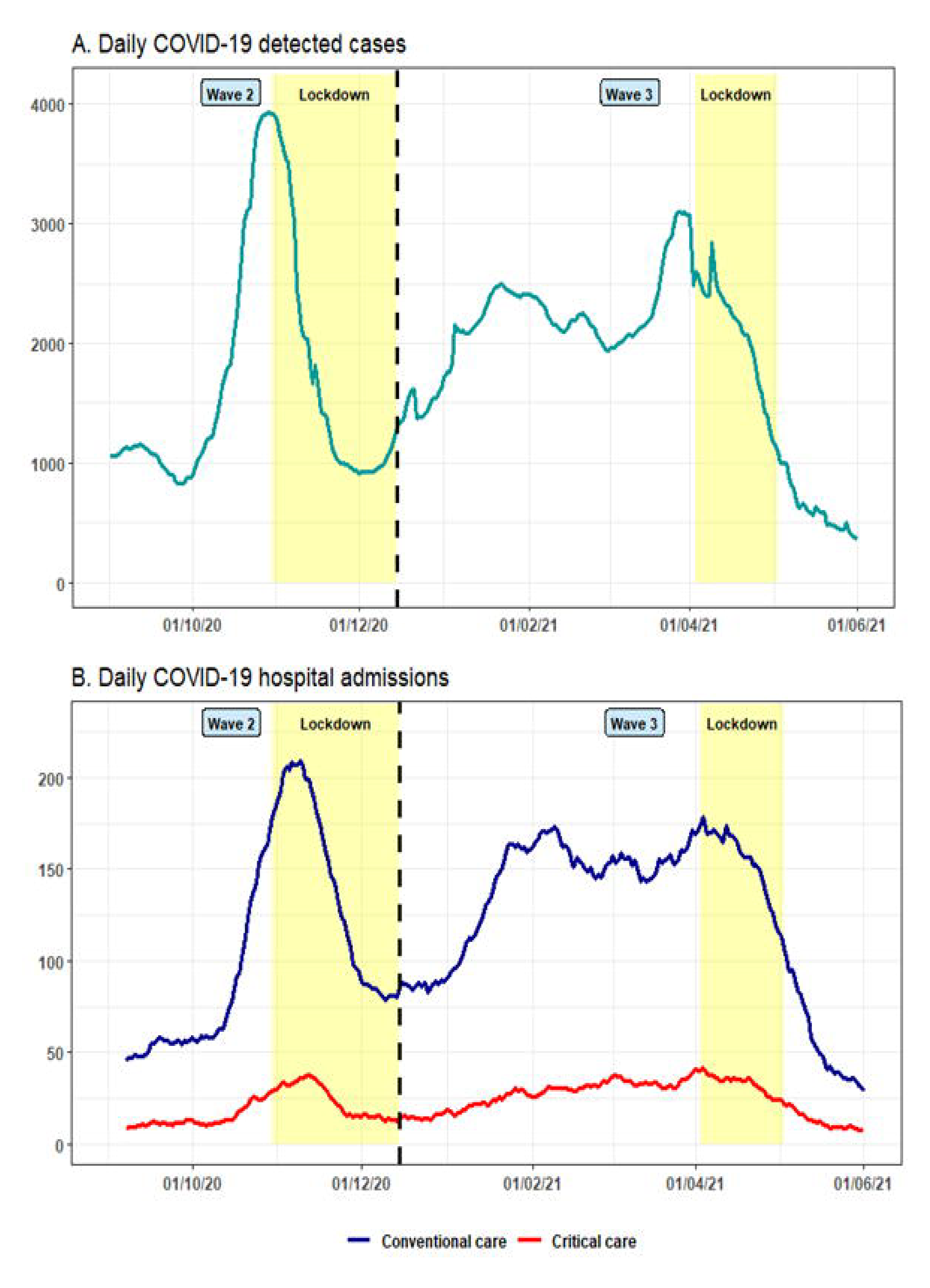
Time evolution of daily COVID-19 cases and hospital admissions, in PACA region, from September 2020 to June 2021. The cutoff between both waves was set at the end of the second French lockdown, which corresponded to a low point in hospital occupancy by COVID-19 patients.

Maps of incidence rates per IRIS (Fig 2, Panel 1) show that the east of the region was less affected during second wave but not during the third one while both waves strongly affected Marseille.

**Figure 2.**
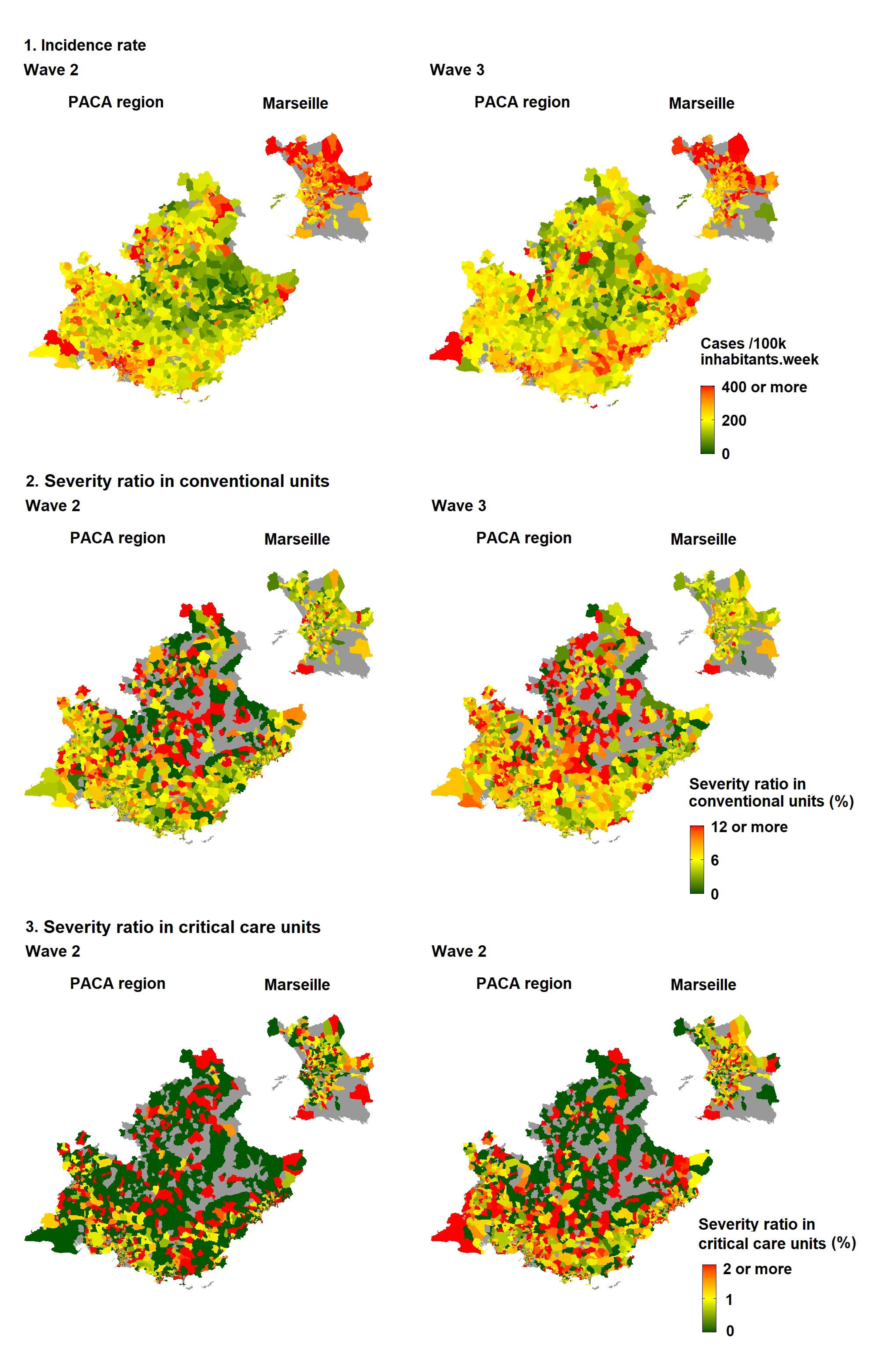
Maps of the incidence rate of COVID-19 cases (panel 1), conventional units severity ratio (panel 2), and critical care units severity ratio (panel 3). Each one at the IRIS level, in PACA region and its main city Marseille, during the second and third epidemic waves.

Moran’s statistics showed a significant, but low, spatial autocorrelation of the incidence rate during the two periods: respectively (resp.) 0.29 (p<0.001) and 0.41 (p<0.001) (Table 1).

**Table 1.** Moran’s index on COVID-19 indicators. This parameter measures the spatial autocorrelation of the indicators between IRIS. It may be interpreted as a classical correlation coefficient. Incidence rate seems to be the indicator most affected by this effect. Wave corresponds to the epidemic wave that hit PACA region, p = p-value.

| Indicator | Wave | Moran's Index | p |
| --- | --- | --- | --- |
| Incidence rates | 2 | 0.29 | < 0.001 |
|  | 3 | 0.41 | < 0.001 |
| Severity ratios in conventional units | 2 | 0.02 | 0.004 |
|  | 3 | 0 | 0.54 |
| Severity ratios in critical care units | 2 | 0 | 0.551 |
|  | 3 | 0.03 | 0.008 |

The temporal evolution of hospital admissions followed that of cases with a small delay (Fig 1, panel B). The severity ratio in conventional units was 6.1% [6.0 - 6.2] and 6.7% [6.7 - 6.8] during the second and third waves, respectively.

This ratio in critical care units was 1.1% [1.0 - 1.1] and 1.4% [1.3 - 1.4] during the second and third waves, respectively.

The two indicators displayed a marked spatial heterogeneity (Fig 2, Panel 2 and 3) and a near-zero spatial autocorrelation index for both waves.

### Description of COVID-19 determinants

We defined six SES profiles which distinguished IRIS on two relevant components: deprivation level and population density (Appendix 2, panels 1.A-1.B). The three middle-income profiles showed very different population densities, which allowed them to be distinguished into “Remote”, “Intermediate” and “Downtown” density profiles. The “Very deprived” IRIS were the least numerous (6.4 %), while “Intermediate” were the most (33.5 %; Appendix 2, panel 1.C). However most deprived IRIS were also very dense, which illustrates the concentration of the most deprived populations in certain areas. These profiles were mapped for the PACA region and its main city, Marseille (Appendix 2, panel 2). At the level of the region, most IRIS covering the largest territory had privileged profiles and were not highly dense. The Marseille urban area was more contrasted with an increasing deprivation gradient from South to North.

We then defined four age structure profiles. Three of them were distinguished by a higher proportion on age category compared to the other profiles, and the fourth presented a balanced distribution (Appendix 2, panels 1.A to 1.D). Thus, “young adults” IRIS had a high proportion of 18-39 years inhabitants, “families” IRIS a high children proportion and a slightly high 18-39 proportion (but less than “young adults” areas), and finally “elderly” IRIS were characterized by a high proportion of elderly people. “Balanced” IRIS, with approximately as many people of each age category, were the most frequent profile (42.8 %) and covered the largest territory in the region (Appendix 2, panel 2). In Marseille, the “families” profile was predominant in the North, and profile with a high proportion of young adults was predominant in the city center.

The mapping of variables characterizing health services provision showed that people living in the mountainous areas in the north and east of the region had less access to frontline caregivers (Appendix 4, panel B). The distance between a given IRIS and the nearest Emergency reception service was more equally distributed: it was less than 50 km in almost the entire region except for a small area in the east (Appendix 4, panel A). The repartition of IRIS with the least healthcare provision seemed to correspond to that of low-density profile areas.

### Explicative models

#### COVID-19 incidence rates

were not associated with SES profile during the second wave, and a slight increase was observed for ”deprived“ IRIS versus “privileged” during the third one (standardized incidence ratio (SIR) of 1.06 CI95% [1.02 – 1.11]; Fig 3, Panels A and B). The age structure seemed to have a bigger impact during both waves: IRIS with a large young adults population reported less cases than other age profiles, for every wave (SIR from 1.06 [1.01 – 1.10] to 1.12 [1.08 – 1.17], with “young adults” profile as a reference class).

**Figure 3.**
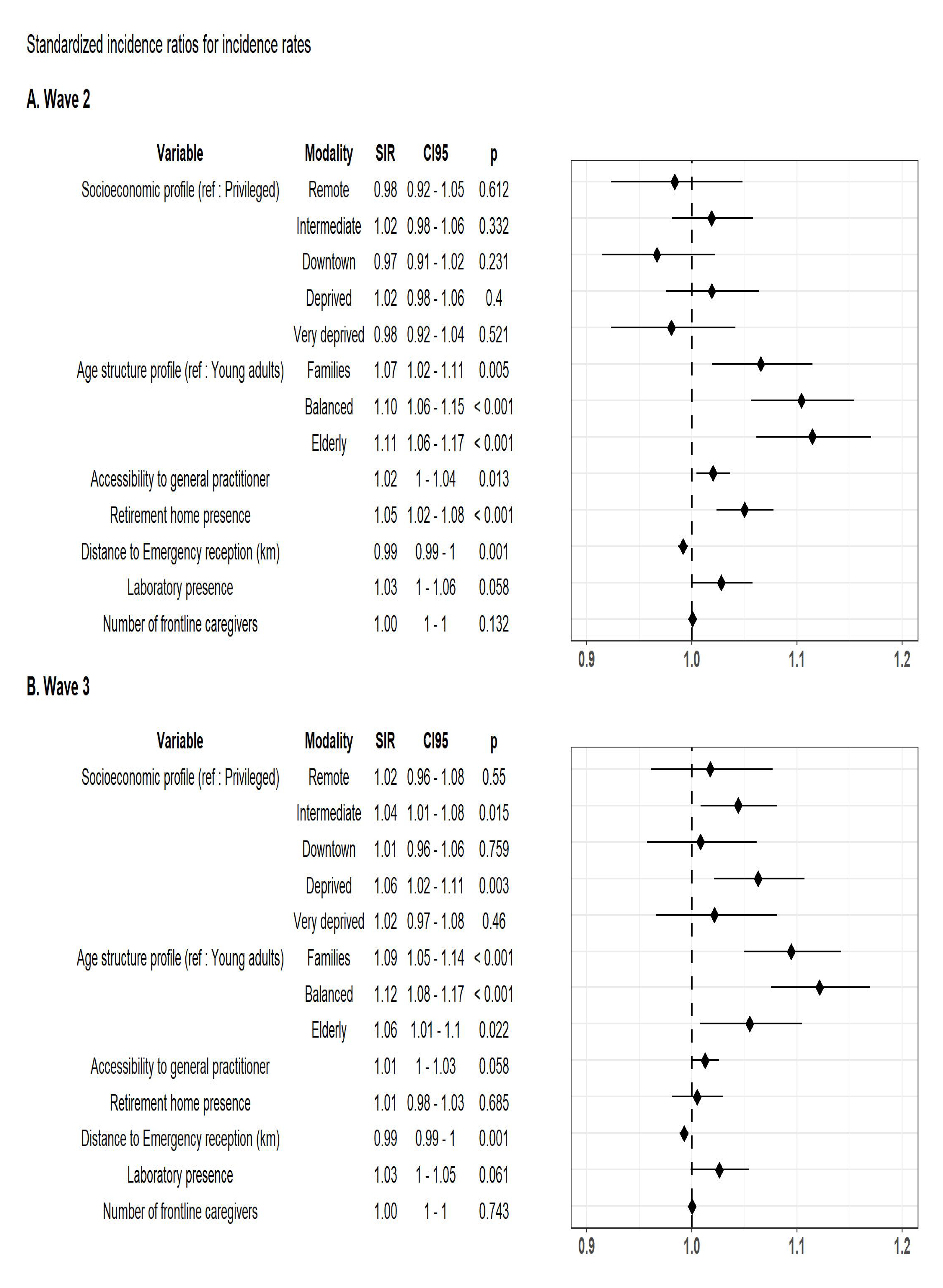
Determinants associated with COVID-19 <u>incidence rates</u> during the second (Panel A) and third (Panel B) epidemic waves, in PACA region, at the IRIS level. SIR = Standardized incidence ratio, CI95 = 95% Confidence Interval, p = p-value.

Furthermore, general practitioners’ accessibility and proximity of an emergency unit were positively associated (resp. p = 0.013 and 0.001) with incidence rates during the second wave. This impact seems to have lessened during the third wave. The impact of the presence of a retirement home seemed to have fallen too between the second and the third waves (SIR from 1.05 [1.02 – 1.08] to 1.01 [0.98 – 1.03]).

#### The severity ratio in conventional units

(conventional-hospitalizations-to-cases ratio), during the second wave higher in IRIS with a “very deprived” than in a “privileged” profile (SIR 1.34 [1.18 - 1.52]) (Fig 4, Panel A). For the “deprived” profile, SIR was 1.25 [1.14 - 1.37]. We also observed this increase in deprived areas during the third wave (SIR 1.25 [1.13 - 1.38] and 1.19 [1.11 - 1.27] for “very deprived” and “deprived” income profiles, respectively) (Fig 4, Panel B). In IRIS with a high proportion of elderly people, this severity ratio increase was greater than 20%, as compared to IRIS with a high proportion of young adults, during both waves (SIR 1.24 [1.11 - 1.38] and 1.22 [1.13 - 1.32], respectively). It was around 10% in IRIS with a retirement home (SIR resp. 1.10 [1.04 - 1.13] and 1.08 [1.04 - 1.13]). The variables characterizing health services provision did not impact these ratios.

**Figure 4.**
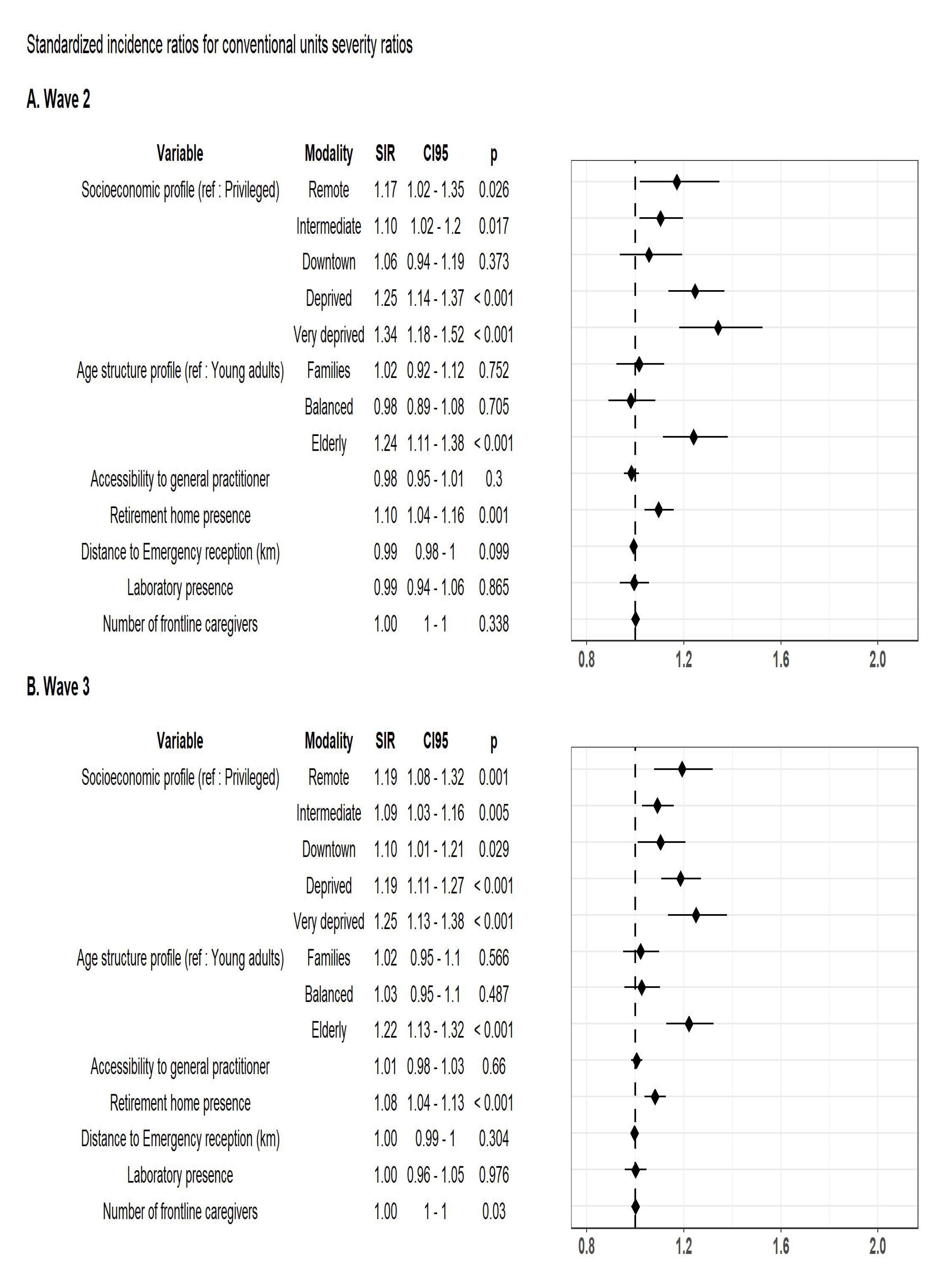
Determinants associated with COVID-19 severity ratios in conventional units (hospitalizations-to-cases ratio) during the second (Panel A) and third (Panel B) epidemic waves, in PACA region, at the IRIS level. SIR = Standardized incidence ratio, CI95 = 95% Confidence Interval, p = p-value

#### The severity ratios in critical care units (critical-care-admissions-to-cases ratios)

for “very deprived” IRIS was even higher than for conventional units (SIR 1.64 [1.30 - 2.07] and 1.33 [1.14 - 1.55] for the second and third wave, respectively, with “privileged” profile as a reference class) (Fig 5, Panels A and B). For ”deprived” IRIS, the gap fades between the second and the third waves (SIR 1.26 [1.05 - 1.50] and 1.00 [0.89 - 1.12], respectively). As in conventional units, IRIS with a large elderly population experienced an increase of around 20% during both waves, although it was not statistically significant during the second one (SIR 1.20 [0.97 - 1.49] and 1.25 [1.09 - 1.44], respectively). None of both periods showed any increase in the IRIS associated with a retirement home. As for conventional care units, this severity ratio was also not associated with variables characterizing access to care.

**Figure 5.**
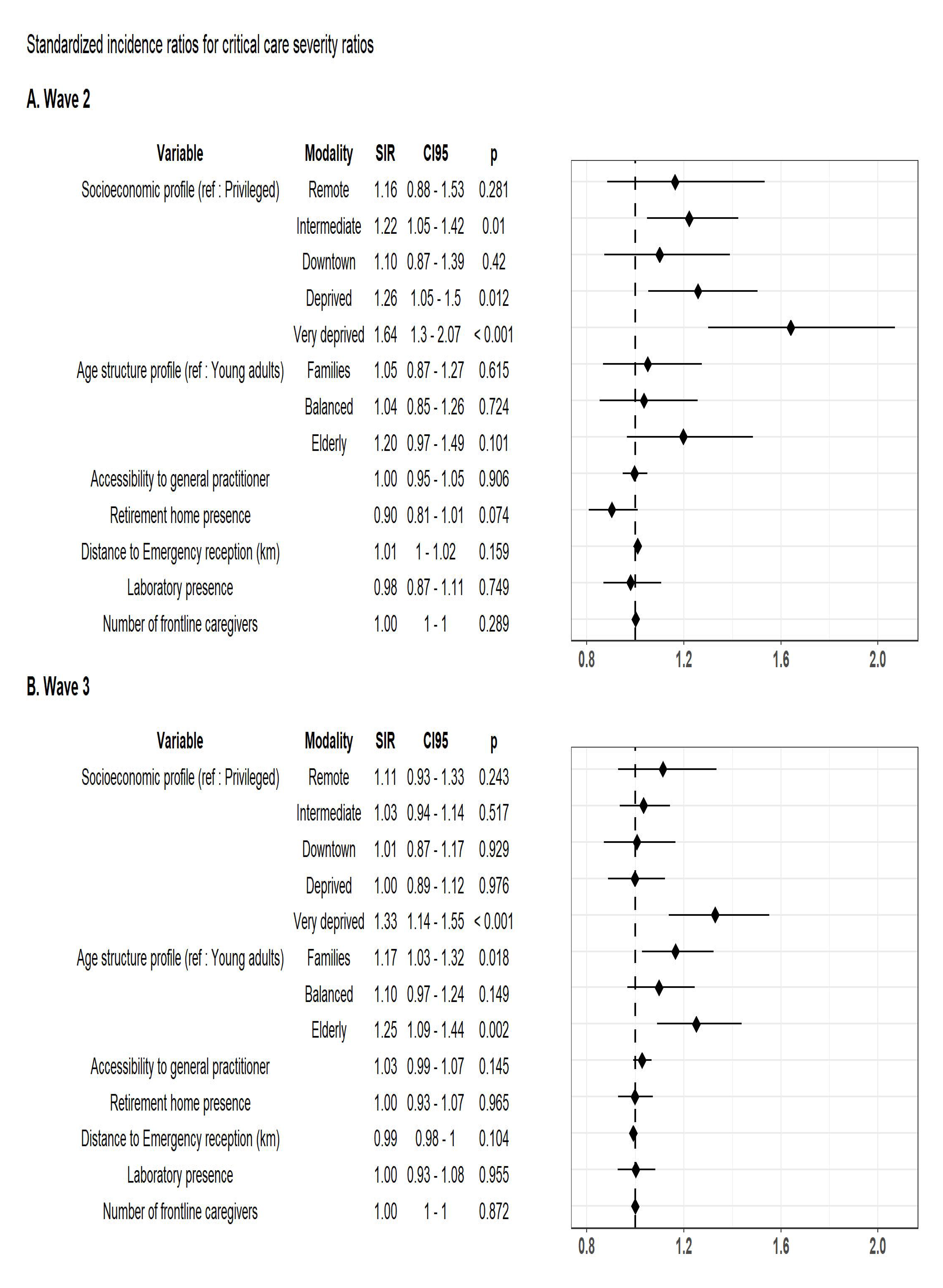
Determinants associated with COVID-19 severity ratios in critical care units (hospitalizations-to-cases ratios) during the second (Panel A) and third (Panel B) epidemic wave, in PACA region, at the IRIS level. SIR = Standardized incidence ratio, CI95 = 95% Confidence Interval, p = p-value

## Discussion

This ecological study showed at a fine geographical local scale that the severity of COVID-19 cases increased in the more deprived IRIS.

Indeed, in "very deprived" areas, the proportion of cases admitted to conventional care units increased during the second pandemic wave by one third compared to "privileged" areas, and by two thirds in critical care. This increase was also observed, at a lower level, in “deprived” areas. This dose-effect relationship argues for the robustness of an effect of SES profile. This agrees with the data from the literature, mainly American and British, analyzed by Wachtler *et al* in their meta-analysis^26^.

There does not seem to be a link between lower access to care and higher severity. Indeed, the variables that characterize it were not associated with a change in the severity of the cases, in either of the periods studied. Other studies do not find a consistent link between COVID-19 severity or incidence indicators and health services provision. Thus, Magesh *et al* observed in the United States that the direction of association with clinical care variables depended on ethnicity^27^.

However, the variables that we considered only reflect the availability of caregivers and services; while lower access to health services could also consist, in deprived populations, of a lower use of care. Indeed, theses populations face various and complex barriers such as discrimination, low literacy, languages, cultural barriers, competing needs and priorities^28, 29^.

Another plausible explanation is the higher prevalence of chronic pathologies in deprived populations, comorbidities being a known determinant of severity linked to COVID-19^30^.

It was notable that the effect of SES was lower during the third pandemic wave compared to the second one. The regional health authority started to implement in early 2021 a mediation action on COVID-19 to reduce social health inequities, with prioritization based on epidemic criteria^31, 32^. This could partly explain, in addition to the COVID-19 global improvement of knowledge, and the first effects of vaccination initially reserved for people at risk, the narrowing of the gap.

The effect of the age structure was, for its part, constant during the two periods considered. The IRIS with the most elderly people had an admission rate between 20 and 25% higher than those with a high proportion of young adults. This result was not surprising since the relationship between advanced age and COVID-19 cases severity is already described^30^.

The presence of a retirement home in the IRIS also had a constant effect during the two waves: an increase in the proportion of admissions around 10% in conventional care, but none in critical care. This apparent paradox could be explained by a lower rate of ICU admission of too elderly and fragile persons^33^. However, it remains difficult to distinguish the legitimate medical refusal of futile invasive treatment, from the triage of eligible patients, in a period of saturation of intensive care services.

We could note during the third wave a strong increase (17%) in the rate of admission to critical care in the IRIS with an age structure of "familial" profile (high proportion of children and young adults, few retirees). It would be relevant to further investigate the potential causes of this increase in severity, not observed in conventional care units.

Finally, no impact of deprivation was observed for incidence during the second wave. This could be linked to greater immunization during the first wave in more deprived IRIS^34^, to a lower use of screening tests due to less perception of hazard^3, 35^, or to a more powerful effect on severity rather than epidemic spread. In the US, Dalton *et al* also did not find an effect of income on test positivity^36^.

One of the limitations of our study was the unavailability of certain determinants that could explain the differences in severity between IRIS. Among them, the proportion of cases of patients with diabetes, obesity, or Chronic Obstructive Pulmonary Disease would be relevant, because of the greater risk of severity described on individual data^16^. As these conditions are more frequent in deprived populations, their effect is, likely, partly included in that attributed to the SES profile.

Another determinant not available at a fine geographical scale is the anti-COVID-19 vaccination rate. Several studies showed greater vaccination of the wealthiest populations^37^, including in France^38^. Vaccination began in France at the beginning of 2021, which corresponds approximately to the start of the third wave, first targeting people with the most fragile health. The COVID-19 severity gap depending on the SES profile was reduced during this wave compared to the second one, so the effect of vaccination, if it existed in our study, would be marginal.

Data regarding the proportion of different ethnicities in the IRIS were also not available. However, data on the proportion of immigrants (foreigners born abroad and residing in France) and foreigners were used to construct SES profiles; they were more numerous in the deprived IRIS. An increase in the risk of severity linked to a minority ethnic group in France could therefore be contained in the results about the SES profile. Indeed, the proportion of black residents seems to be the SES determinant most strongly associated to COVID-19 fatalities^7, 10^ and seroprevalence^34^.

The two variables representing IRIS profiles, those of SES and age structure, were collinear. One risk of this collinearity was the instability of the coefficients. This could explain, for instance, the isolated increase in the critical care admission rate in “familial” IRIS during the third wave. It was simultaneous with a clear decrease in the “low-income” profiles, and more than a third of “family” profile IRIS are also “low-income” profile ones. The robustness of the other results through several analysis models nevertheless suggests that this collinearity had otherwise little influence on the estimated coefficients.

The addresses were declarative and carried a risk of inaccuracy, but the very large number of data compensated for this. The IRIS were also sometimes heterogeneous, with for example slums in the central districts of Marseille.

Finally, the spatial analysis well considered the autocorrelation between neighboring IRIS as shown by the null Moran coefficients on the residuals of our models (Table 2).

**Table 2.** Moran’s index on GAMM residual models for the different considered indicators. Where the spatial autocorrelation was strong between indicators, it was weak or nill between models’ residuals.

| Indicator | Wave | Moran's Index | p |
| --- | --- | --- | --- |
| Incidence rates | 2 | 0.02 | 0.1 |
|  | 3 | 0.03 | 0.014 |
| Severity ratios in conventional units | 2 | -0.04 | 0.996 |
|  | 3 | 0.03 | 0.014 |
| Severity ratios in critical care units | 2 | -0.02 | 0.875 |
|  | 3 | -0.01 | 0.788 |

In conclusion, this ecological study has shown, on a fine geographical scale, that deprivation was associated with greater severity of COVID-19 cases.

This result implies targeting risk prevention efforts on these areas in pandemic situations. Anti-COVID mediation teams have been set up for this purpose, and the benefits of their action are being assessed. These results also call for more studies at the individual level to understand the determinants involved in this increase in hospital admissions.

## Statements

### Ethics approval

To ensure the confidentiality of the information and in accordance with French regulations, only data aggregated to the IRIS and to the day were processed for this study. Clearance was obtained through a specific convention (number 22DIRA41-0) between Aix Marseille University and Santé publique France, from the Aix Marseille University Ethic committee (number 2022-10-20-006), and from the Aix Marseille University Data Protection Officer (number 513087).

### Patient and public involvement

The public was not involved in the preparation or conduct of this study, but dissemination of its results is planned both to policy makers and to organizations called upon to support prevention actions.

## Data Availability

All data produced in the present study are available upon reasonable request to the authors

## Acknowledgement

Agence régionale de santé PACA, Santé Publique France

## Data sharing

Data collected and aggregated by Santé Publique France and Agence régionale de santé PACA.

