## Appendix for "Deprivation effect on COVID-19 cases incidence and severity: a geo-epidemiological study in PACA region, France"

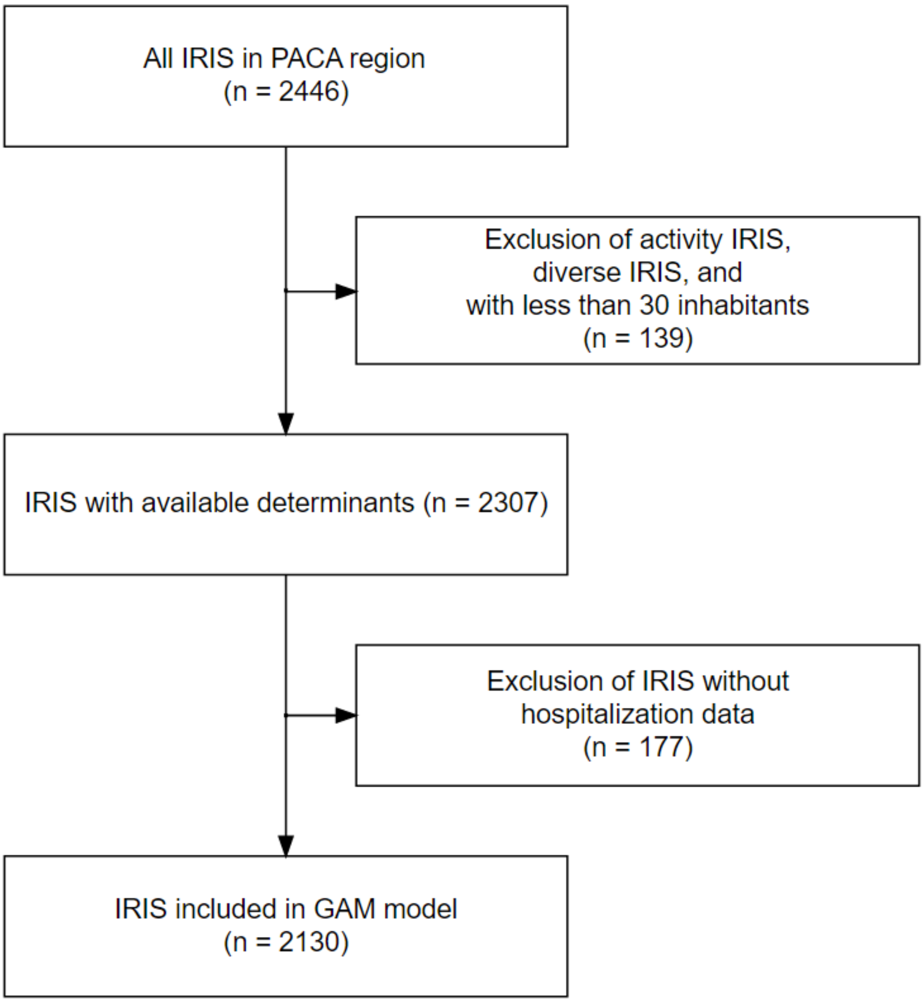


**Appendix 1. Flow chart of IRIS selection.**

2130 IRIS (4.92 million inhabitants) were included among the 2446 IRIS in PACA region (5.04 million inhabitants).

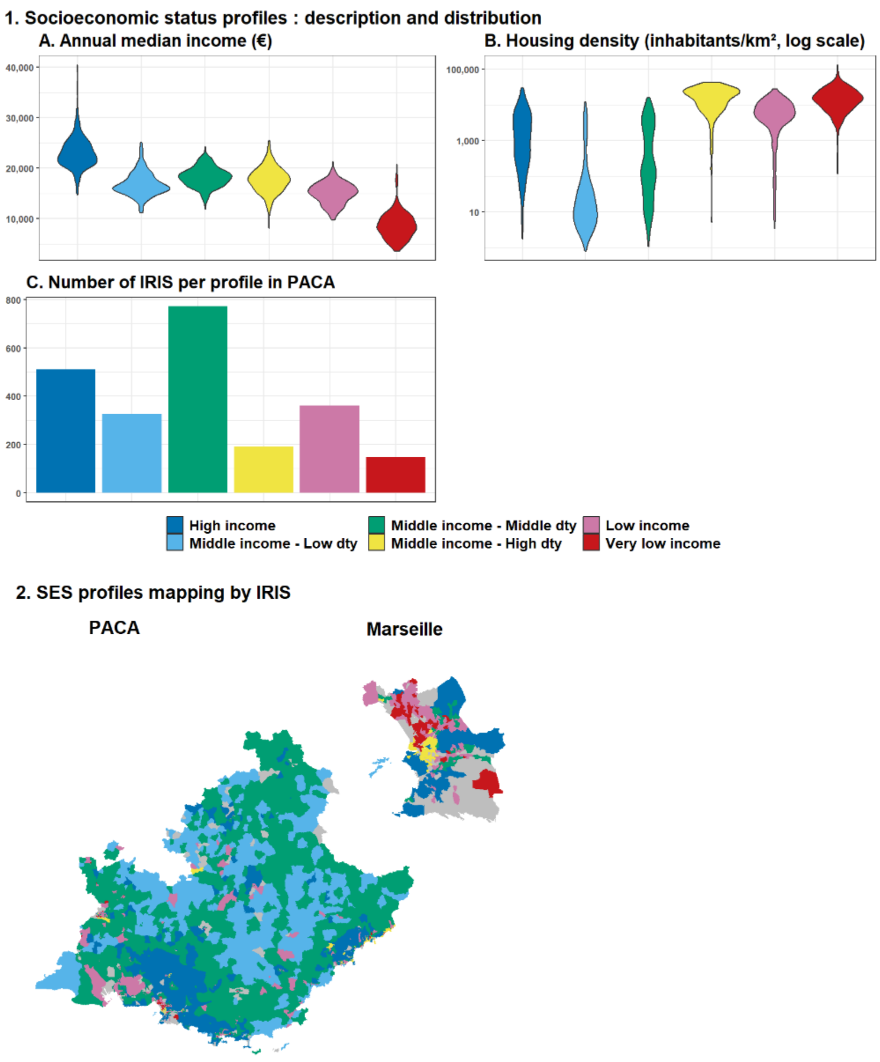


**Appendix 2.**

**1. Distribution of annual median income and population density in each socioeconomic status profile (Panels A-B) in PACA, and number of IRIS in each one (Panel C).** Profiles were defined by unsupervised Hierarchical classification on Principal Components (HCPC), using SES factors available on the national INSEE website. The names used for the classes sum up the class components. Three profiles had similar income ("middle income") distributions but differed strongly on population density (dty). Most deprived IRIS were the most densely populated.

**2. Map of the socioeconomic status profiles, at the IRIS level in PACA region**

The color scale represents the different profiles. The NA (grey) class corresponds to unpopulated areas.

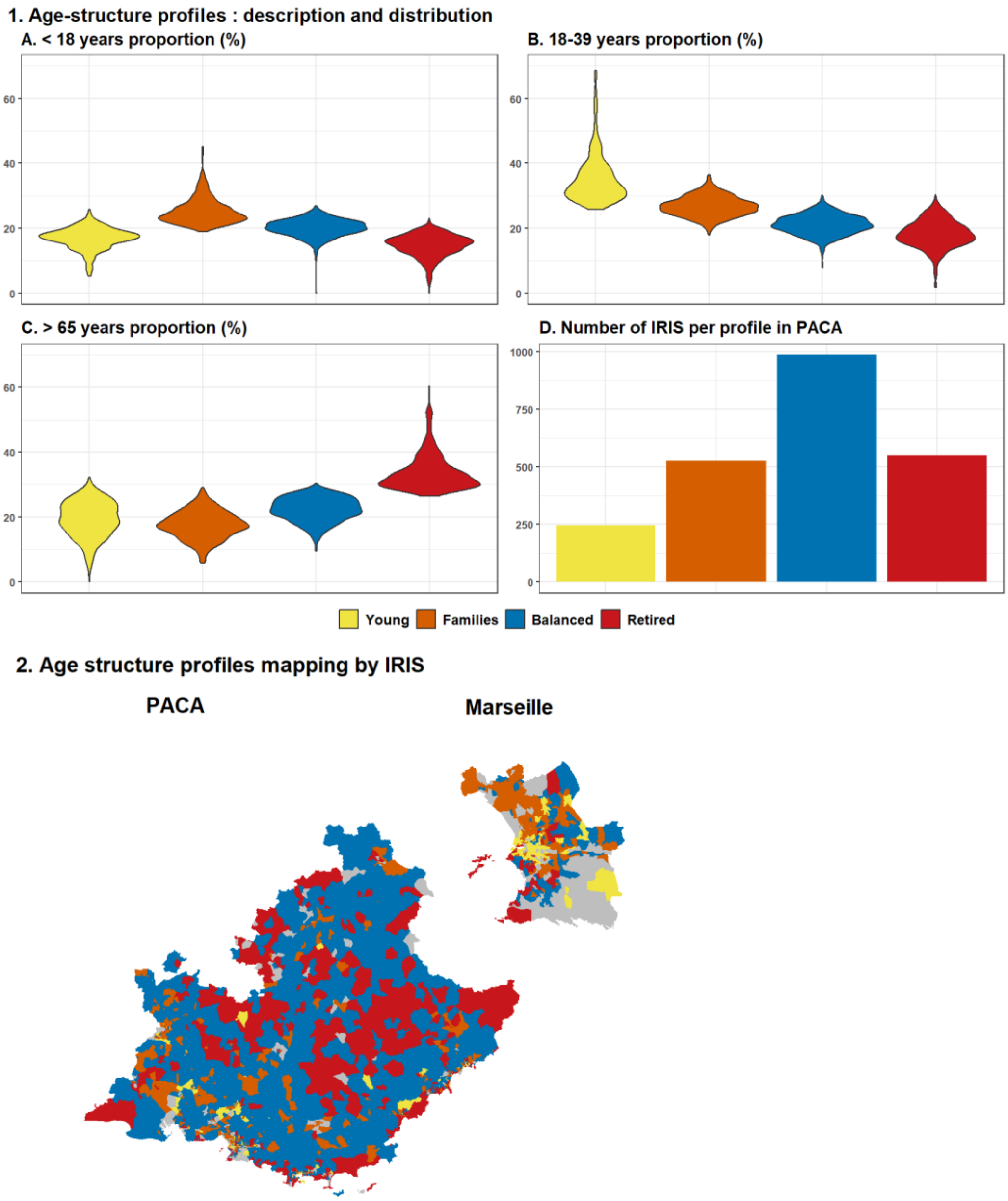


**Appendix 3.**

**1. Distribution of age category proportions in each age structure profile (panels A-C) in PACA, and number of each one (panel D).** Proportions of each profile defined by HCPC allow to define four age-structure profiles. "Families"-structure profile had a lot of children, "young" profile had a great proportion of 18-39 years inhabitants. A well-balanced class was observed, it corresponded to most of the IRIS.

**2. Map of the age-structure profiles, at the IRIS level in PACA region and its main city Marseille.** The color scale represents the different profiles. The NA (grey) class correspond to unpopulated areas. "Balanced"-structured profile IRIS covered most of the territory outside the cities.

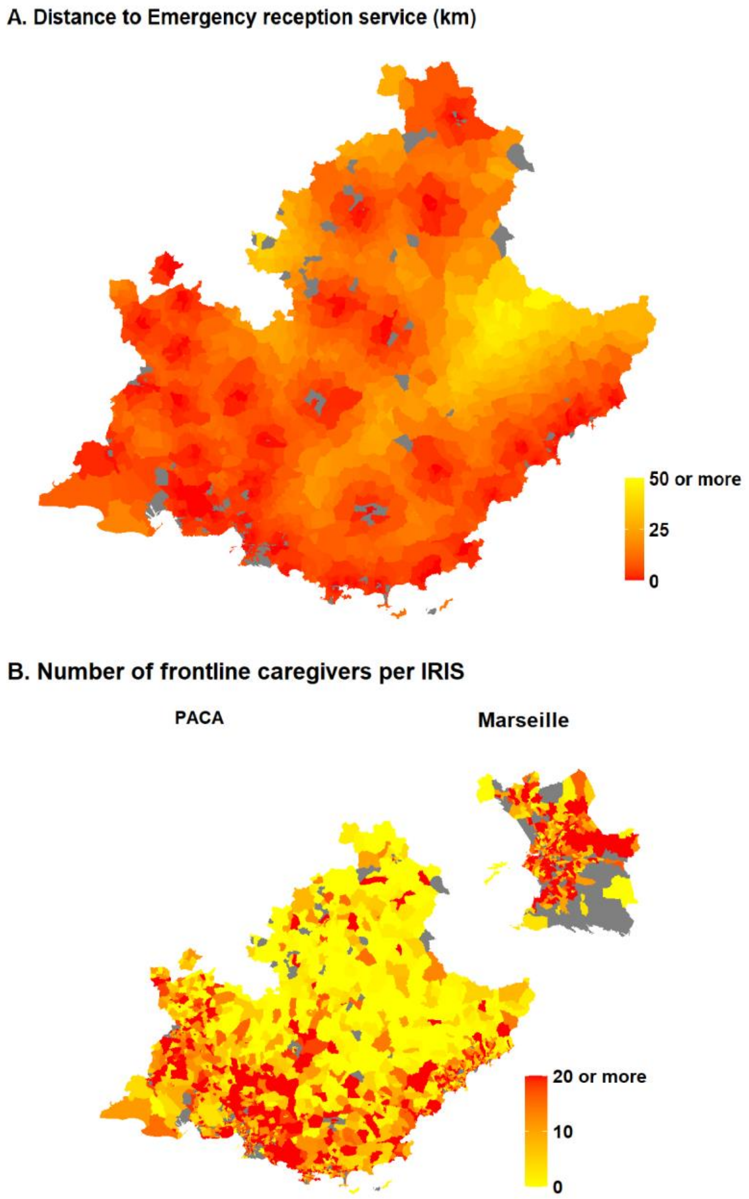


**Appendix 4.**

**Panel A : Map of the distance to Emergency reception service for each service, at the IRIS level in PACA region.** The NA (grey) class correspond to unpopulated areas. Distances were calculated between IRIS centroids. The IRIS centroids of ones with an emergency reception service served as base points for the calculation. There was at least one service in each big city.

**Panel B : Map of the number of frontline caregivers per IRIS, in PACA region and its main city Marseille.** The NA (grey) class correspond to unpopulated areas. List of frontline caregivers (INSEE): nurses, general practitioners, gynecologists, pediatricians, psychiatrists, ophthalmologists, dental surgeons, midwives, physiotherapists.

**Appendix 5. List of frontline caregivers** (i.e., primary care providers, and other care professionals directly accessible for the patients, as defined by Department of Research, Studies, Evaluation and Statistics): nurses, general practitioners, gynecologists, pediatricians, ophthalmologists, psychiatrists, dental surgeons, midwives, physiotherapists.

### Appendix 6. Database sources

| Database name | Date of publication | Last access to the database | Author | Link |
|---|---|---|---|---|
| **Equipment** | 12/07/2021 | 13/07/2021 | INSEE | https://www.insee.fr/fr/statistiques/3568629?sommaire=3568656&q=bpe+2020 |
| **Census** | 09/12/2020 | 12/04/2021 | INSEE | https://www.insee.fr/fr/statistiques/4515565?sommaire=4516122&q=recensement+2017#consulter |
| **Housing** | 09/12/2020 | 21/05/2021 | INSEE | https://www.insee.fr/fr/statistiques/4515532?sommaire=4516107&q=base+logement#dictionnaire |
| **EDI 2015** | 01/01/2015 | 21/05/2021 | MapInMed | https://www.anticipe.eu/plateformes/MAPinMED |
| **IRIS borders (shapefile)** | | 21/05/2021 | Data.gouv | https://www.data.gouv.fr/en/datasets/decoupage-iris-combine-aux-limites-communales-openstreetmap/ |
| **Accessibility to general practitioners** | 02/03/2020 | 30/07/2021 | DREES | https://drees2-sgsocialgouv.opendatasoft.com/explore/dataset/530_l-accessibilite-potentielle-localisee-apl/information/ |

### Appendix 7. List of variables included in HCPC

List of variables included in HCPC to define SES profiles :

- Habitat density
- Proportions of each socio-professional category:
  - Traders
  - White collar workers
  - Intermediate professions
  - Employees
  - Blue collar workers
  - Retired
  - Farmers
  - Without professional activity
- Proportion of foreigners
- Proportion of immigrants
- Proportion of bachelor
- Median income
- Proportion of overcrowded housing
- European Deprivation Index

List of variables included in HCPC to define age structure profiles:

- Proportion of 0–17-year-olds
- Proportion of 18–39-year-olds
- Proportion of 40–64-year-olds
- Proportion of people over 65
